# Long-term benefits of breastfeeding on brain and body development among 9–10-year-olds: modulated by socioeconomic environment

**DOI:** 10.1101/2023.01.06.23284287

**Authors:** Vidya Rajagopalan, Eustace Hsu, Shan Luo

## Abstract

**Importance:** It is yet unknown if breastfeeding (bf) benefits, to brain and body development of children, persist into peri-adolescence and vary by socioeconomic environments (SEEs).

**Objective:** We aim to investigate SEE-independent and SEE-modulated relationships between bf duration and child brain structure and adiposity markers during peri-adolescence.

**Design, setting and participants:** This was a cross-sectional study of children aged 9–10 enrolled in the multi-center Adolescent Brain and Cognitive Development (ABCD) Study®.

**Exposure(s):** Bf duration was self-reported. Neighborhood-level SEE was assessed using area deprivation index (ADI).

**Main Outcome(s):** T1-weighted magnetic resonance imaging was used to assess global brain measures: volumes of white, cortical, and subcortical gray matter (GM), cortical thickness, and surface area (SA). Adiposity markers included age- and sex-specific body mass index (BMI *z-*scores), waist circumference, and waist-to-height ratio (WHtR). Mixed effects models examined associations of bf duration with brain structure and adiposity markers controlling for sociodemographic, pre- and post-natal covariates. Stratified analysis was performed by tertiles of ADI.

**Results:** The sample consisted of 7,511 children (51.7% males; 18.8% no bf, 35.3% 1-6 months, 24.9% 7-12 months, 21.0% >12 months). Child’s total SA (β (95% CI) = 0.053 (0.033, 0.074); FDR corrected *P*<0.001), cortical (β (95% CI) = 0.021 (0.010, 0.032); FDR corrected *P*<0.001) and subcortical GM volume (β (95% CI) = 0.016 (0.003, 0.030); FDR corrected *P*<0.001) increased monotonically with bf duration, after controlling for covariates. Child’s BMI *z*-scores (β (95% CI) = -0.040 (−0.063, -0.016); FDR corrected *P*=0.001), waist circumference (β (95% CI) = -0.037 (−0.060, -0.014), FDR corrected *P*=0.002) and WHtR (β (95% CI) = -0.040 (−0.064, -0.018), FDR corrected *P*=0.001) decreased monotonically with increased bf duration, after controlling for covariates. Bf duration was inversely associated with adiposity in children from high- and medium-ADI neighborhoods. Bf duration was positively associated with SA across ADI tertiles.

**Conclusions and Relevance:** Our results imply that long-term benefits of bf on body and brain development in offspring increase as bf duration increases, particularly in children from low SEEs. Policies and social support aimed to incremental increases in bf duration among women from low SEEs would confer long-term benefits for offspring.

**Key Points:** *Question:* Do benefits of breastfeeding(bf), on children’s brain and body development, persist long-term and are these benefits uniform across socioeconomic environments (SEEs)?

*Findings:* Longer bf duration is associated with lower adiposity, greater cortical and subcortical gray matter volume, and cortical surface area in 9–10-year-old children. Children from lower SEEs showed stronger negative relationships between bf duration and adiposity. Children across all SEEs demonstrated positive relationships between bf duration and surface area.

*Meaning:* Our results imply that long-term benefits to child brain and body development increase with bf duration; and children from lower SEEs benefited more from longer bf duration.

## Introduction

The short-term, multi-faceted benefits of breastfeeding (bf) for offspring brain and body development have been well-established. It is known that longer bf duration correlates with healthy weight gain and lower rates of infant obesity^1^. Additionally, multiple studies have shown that longer bf duration promotes better brain white matter (WM) development in infancy, which is associated with better childhood neurodevelopmental outcomes^2,3^. But there is limited knowledge on if these benefits persist into late childhood^4^.

Current knowledge of the benefits of bf on brain and body development in youth are limited using proxies such as obesity incidence and school performance. In addition to not being informative about underlying biological processes, these proxies are also strongly influenced by socioeconomic factors. Socioeconomic environment (SEE) is an important influencer of childhood development with higher SEE levels correlated with lower adiposity and greater brain structural measures^5–7^. Furthermore, studies tend to overstate long-term benefits of bf because of selection bias towards high SEE families who are more likely to breastfeed longer^8–10^. It remains unclear if long-term benefits of bf exist after controlling for SEE levels and if these benefits vary by SEEs.

In this study, we leveraged demographic, gestational and infant health history, anthropometric and brain data from the Adolescent Brain Cognitive Development (ABCD) study, the largest long-term study of pediatric brain development in the U.S., to determine relationships of bf duration with brain structure and adiposity markers in a cohort of 7511 9–10-year-olds from various SEE levels. Using these measures, as opposed to obesity incidence or test scores, we can better investigate the underlying biological changes associated with bf. We hypothesize that length of bf will have an inverse relationship with adiposity measures and positive relationship with structural brain measures independent of SEE levels. Based on prior work, we anticipate that the benefits of bf will be more pronounced in the higher SEE groups^11,12^.

## Methods

### Participants

Data was obtained from the baseline assessments of ABCD 3.0 data release (N=11,875). Details of ABCD study design, recruitment and inclusion/exclusion were mentioned elsewhere^13^. After applying study-specific exclusion criteria on anthropometric measurements, neuroimaging, and other covariates as described in the next sections, our final sample size is 7511.

### Breastfeeding length

Breastfeeding duration was self-reported by the caregiver and was coded into 4 subcategories: no breastfeeding (0 months), 1-6 months, 7-12 months, >12 months. Categorizations were made with consideration to guidelines from the American Academy of Pediatrics, for synchronization with cut points used elsewhere in the literature, and to ensure adequate sample sizes within subgroupings^14–17^.

### Socioeconomic environment Family level

Parental education and annual family income were used as indexes of family level SEE. Parental education was coded as a binary variable indicating whether at least one parent has obtained a bachelor’s degree. Yearly household income places combined income into three categories: less than $50,000, between $50,000 and $99,999, and at least $100,000.

### Neighborhood level

Area Deprivation Index (ADI) was used to measure neighborhood-level SEE. ADI is a combination of characteristics of a neighborhood reported from the American Community Survey (2011-2015)^18,19^, with higher ADI score representing lower SEE. For this study, we computed ADI from primary addresses that were matched with 2010 US census tracts in order to identify neighborhoods of residence^20^. ADI percentile score was computed for each participant corresponding to their primary residence, then was grouped into tertiles for ease of interpretation^20^.

### Adiposity markers

Weight, height, and waist circumference were recorded by a trained researcher at baseline visit and used to calculate waist-to-height ratio (WHtR) and body mass index (BMI, kg/m^2^).

Measurements were further converted into age- and sex-specific BMI percentiles, BMI *z*-scores, weight *z*-scores, height *z*-scores and waist *z*-scores^21,22^. The following criteria were used to exclude anthropometrics data: 1) BMI *z*-scores ≤ -4 SDs or ≥8 SDs, 2) BMI <10, 3) weight *z*-scores ≤-5 SDs or ≥8 SDs, 4) height *z*-scores or waist *z*-scores <-4 SDs or >4 SDs, 5) WHtR ≤0.2 or ≥1^23,24^.

### Neuroimaging

T1-weighted anatomical scans were collected using methods optimized for 3-T scanners across multiple platforms at 21 ABCD Study sites^25,26^. T1 Images that passed ABCD quality control were segmented using FreeSurfer (version 5.3.0), yielding the following measures: total volumes of cortical gray, subcortical gray (GM) and white matter (WM) (mm^3^), cortical thickness (mm), and cortical surface area (SA) (mm^2^)^27^.

### Other relevant covariates

The following variables were extracted from the ABCD database: race/ethnicity, pubertal stage, gestational age, maternal health problems during pregnancy, child health problems at birth, maternal age, and maternal alcohol or tobacco use during pregnancy. Further details can be found in supplementary methods^28,29^.

### Data Analysis

All analyses were conducted in R. Individual linear mixed effects models were used to examine correlations between bf length (independent variable) and each global brain measurement and adiposity marker (dependent variable) for the whole cohort, while controlling for family-level SEE and other relevant covariates. Age and sex were not included as covariates for models with BMI *z*-scores as a dependent variable. Models of brain measurements included handedness, and intracranial volume for volumetric measures. Family ID, nested within site, were modeled as random intercepts in models of adiposity markers; Family ID, nested within scanner ID, were modeled as random intercepts for models of brain measurements, to account for shared variation related to family membership and study visit location and/or scanner.

We investigated the interaction of ADI and bf duration only on those brain and adiposity measures identified as significant in the whole cohort model, followed by stratified analysis by each ADI tertile.

Linear mixed effects models used the *lme4* package^30^, with *P*-values calculated using Satterthwaite’s method in the *lmerTest* package^31^. Standardized betas were reported. 95% Wald confidence intervals were calculated based on the local curvature of the likelihood surface. Tests of significance (2-tailed) were corrected for multiple comparisons using the Benjamini-Hochberg false discovery rate (FDR) correction, with *P*<0.05 as the corrected threshold for significance.

## Results

### Participant Demographics

**Table 1** describes characteristics of the sample by bf length category. 18.8% of child participants were breastfed for 0 months, 35.3% for 1-6 months, 24.9% for 7-12 months, and 21.0% for greater than 12 months. Participants breastfed for different lengths (by category) differed in terms of age at baseline (*F*(3, 7507) = 6.77, *P*=0.0001), race/ethnicity (X^2^(12) = 586.52, *P*<0.001), family income (X^2^(6) = 493.92, *P*<0.001), parental education (X^2^(3) = 767.79, *P*<0.001), pubertal status (X^2^(6)= 200.79, *P*<0.001), gestational age (*F*(3, 7507) = 67.00, *P*<0.001), maternal health problems (X^2^(3) = 127.84, *P*<0.001), health problems at birth (X^2^(3) = 20.85, *P*=0.0001), maternal age at birth (*F*(3, 7507) = 113.83, *P*<0.001), and maternal alcohol and/or pregnancy use (X^2^ (3)= 28.50, *P*<0.001).

**Table 1.** Sample Characteristics by Breastfeeding Length Category.

|  | No<br>Breastfeeding<br>(N=1412) | 1-6 months<br>(N=2651) | 7-12<br>months<br>(N=1873) | > 12<br>months<br>(N=1575) | Total(N=7511) |  |
| --- | --- | --- | --- | --- | --- | --- |
|  | Mean (SD) or N<br>(%) | Mean<br>(SD) or N<br>(%) | Mean<br>(SD) or N<br>(%) | Mean<br>(SD) or N<br>(%) | Mean (SD) or<br>N (%) | *P*-value<sup>a</sup> |
| Age (years) | 9.928 (0.607) | 9.952<br>(0.628) | 9.882<br>(0.631) | 9.878<br>(0.632) | 9.915<br>(0.626) | **0.0001\*\*\*** |
| Sex |  |  |  |  |  |  |
| Female | 667 (47.2%) | 1307<br>(49.3%) | 897<br>(47.9%) | 759<br>(48.2%) | 3630<br>(48.3%) | 0.608 |
| Male | 745 (52.8%) | 1344<br>(50.7%) | 976<br>(52.1%) | 816<br>(51.8%) | 3881<br>(51.7%) |  |
| Race/Ethnicity |  |  |  |  |  |  |
| White | 606 (42.9%) | 1425<br>(53.8%) | 1212<br>(64.7%) | 1065<br>(67.6%) | 4308<br>(57.4%) | **<0.001\*\*\*** |
| Black | 389 (27.5%) | 318<br>(12.0%) | 114<br>(6.1%) | 56 (3.6%) | 877 (11.7%) |  |
| Hispanic | 263 (18.6%) | 586<br>(22.1%) | 323<br>(17.2%) | 265<br>(16.8%) | 1437<br>(19.1%) |  |
| Asian | 8 (0.6%) | 44 (1.7%) | 38 (2.0%) | 39 (2.5%) | 129 (1.7%) |  |
| Other | 146 (10.3%) | 278<br>(10.5%) | 186<br>(9.9%) | 150<br>(9.5%) | 760 (10.1%) |  |
| Family Income |  |  |  |  |  |  |
| <\$50,000 | 661 (46.8%) | 751<br>(28.3%) | 359<br>(19.2%) | 278<br>(17.7%) | 2049<br>(27.3%) | **<0.001\*\*\*** |
| ≥\$50,000 and<br><\$100,000 | 419 (29.7%) | 754<br>(28.4%) | 519<br>(27.7%) | 478<br>(30.3%) | 2170<br>(28.9%) | |
| ≥\$100,000 | 332 (23.5%) | 1146<br>(43.2%) | 995<br>(53.1%) | 819<br>(52.0%) | 3292<br>(43.8%) | |
| Parental Education |  |  |  |  |  |  |
| No Bachelor's<br>Degree | 900 (63.7%) | 1042<br>(39.3%) | 445<br>(23.8%) | 324<br>(20.6%) | 2711<br>(36.1%) | **<0.001\*\*\*** |
| Bachelor's<br>Degree or Higher | 512 (26.3%) | 1609<br>(60.7%) | 1428<br>(76.2%) | 1251<br>(79.4%) | 4800<br>(63.9%) |  |
| Pubertal Stage |  |  |  |  |  |  |
| Pre-Puberty | 609 (43.1%) | 1281<br>(48.3%) | 1112<br>(59.4%) | 942<br>(59.8%) | 3944<br>(52.5%) | **<0.001\*\*\*** |
| Early Puberty | 312 (22.1%) | 659<br>(24.9%) | 428<br>(22.9%) | 352<br>(22.3%) | 1751<br>(23.3%) |  |
| Mid-Post Puberty | 491 (34.8%) | 711<br>(26.8%) | 333<br>(17.8%) | 281<br>(17.8%) | 1816<br>(24.2%) |  |
| Gestational Age at<br>Birth | 38.832<br>(2.426) | 38.788<br>(2.466) | 38.391<br>(1.725) | 39.586<br>(1.423) | 39.114<br>(2.130) | **<0.001\*\*\*** |
| Maternal Health<br>Problems During<br>Pregnancy |  |  |  |  |  |  |
| No | 760 (53.8%) | 1547<br>(58.4%) | 1277<br>(68.2%) | 1102<br>(70.0%) | 4686<br>(62.4%) | **<0.001\*\*\*** |
| Yes | 652 (46.2%) | 1104<br>(41.6%) | 596<br>(31.8%) | 473<br>(30.0%) | 2825<br>(37.6%) |  |
| Child Health<br>Problems at Birth |  |  |  |  |  |  |
| No | 1046 (74.1%) | 1843<br>(69.5%) | 1375<br>(73.4%) | 1187<br>(75.4%) | 5451<br>(72.6%) | **0.0001\*\*\*** |
| Yes | 366 (25.9%) | 808<br>(30.5%) | 498<br>(26.6%) | 388<br>(24.6%) | 2060<br>(27.4%) |  |
| Mother's Age at<br>Birth | 27.381<br>(6.514) | 29.547<br>(6.219) | 30.482<br>(5.481) | 31.140<br>(5.445) | 29.707<br>(6.077) | **<0.001\*\*\*** |
| Maternal Alcohol<br>or Tobacco use<br>During Pregnancy |  |  |  |  |  |  |
| No | 898 (63.6%) | 1695<br>(63.9%) | 1322<br>(70.6%) | 1070<br>(67.9%) | 4985<br>(66.4%) | **<0.001\*\*\*** |
| Yes | 514 (36.4%) | 956<br>(36.1%) | 551<br>(29.4%) | 505<br>(32.1%) | 2526<br>(33.6%) |  |

<sup>a</sup> \*\*\* $P < 0.001$

### Breastfeeding Length and Child Global Brain Measurements

BF duration was positively associated with total cortical SA (β (95% CI) = 0.053 (0.033, 0.074), FDR corrected *P*<0.0001, **Figure 1A**), cortical (β (95% CI) = 0.021 (0.010, 0.032), FDR corrected *P*=0.0002) and subcortical GM volume (β (95% CI) = 0.016 (0.003, 0.030), FDR corrected *P*=0.025), and negatively associated with WM volume (β (95% CI) = -0.022 (−0.033, -0.011), FDR corrected *P*=0.0003) (**Table 2**, Supplemental Figure 1A).

**Table 2.**
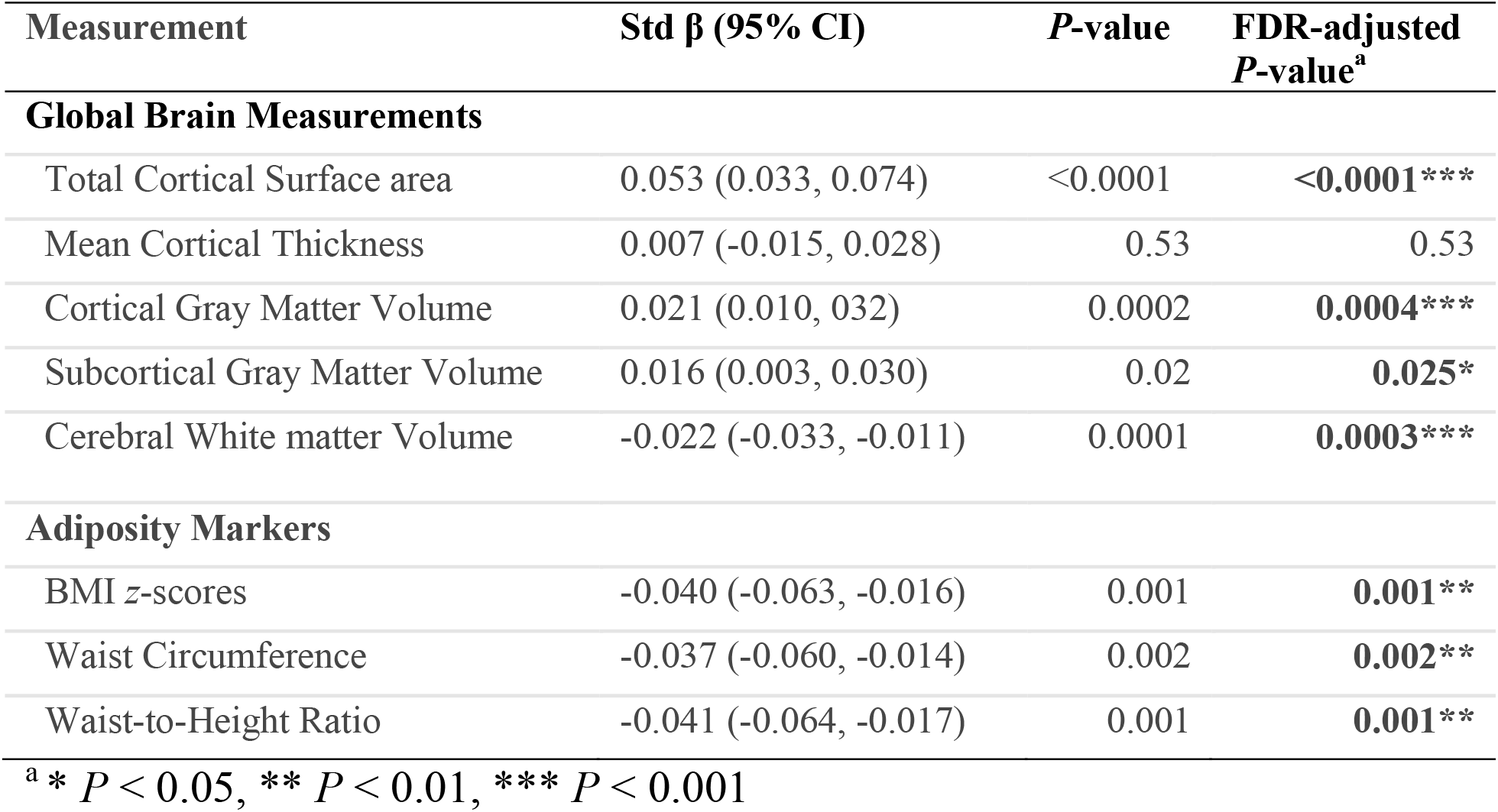
Associations between Breastfeeding Length Category (Monotonic Relationship) and Child Adiposity Markers, Global Brain Measurements.

**Figure 1.**
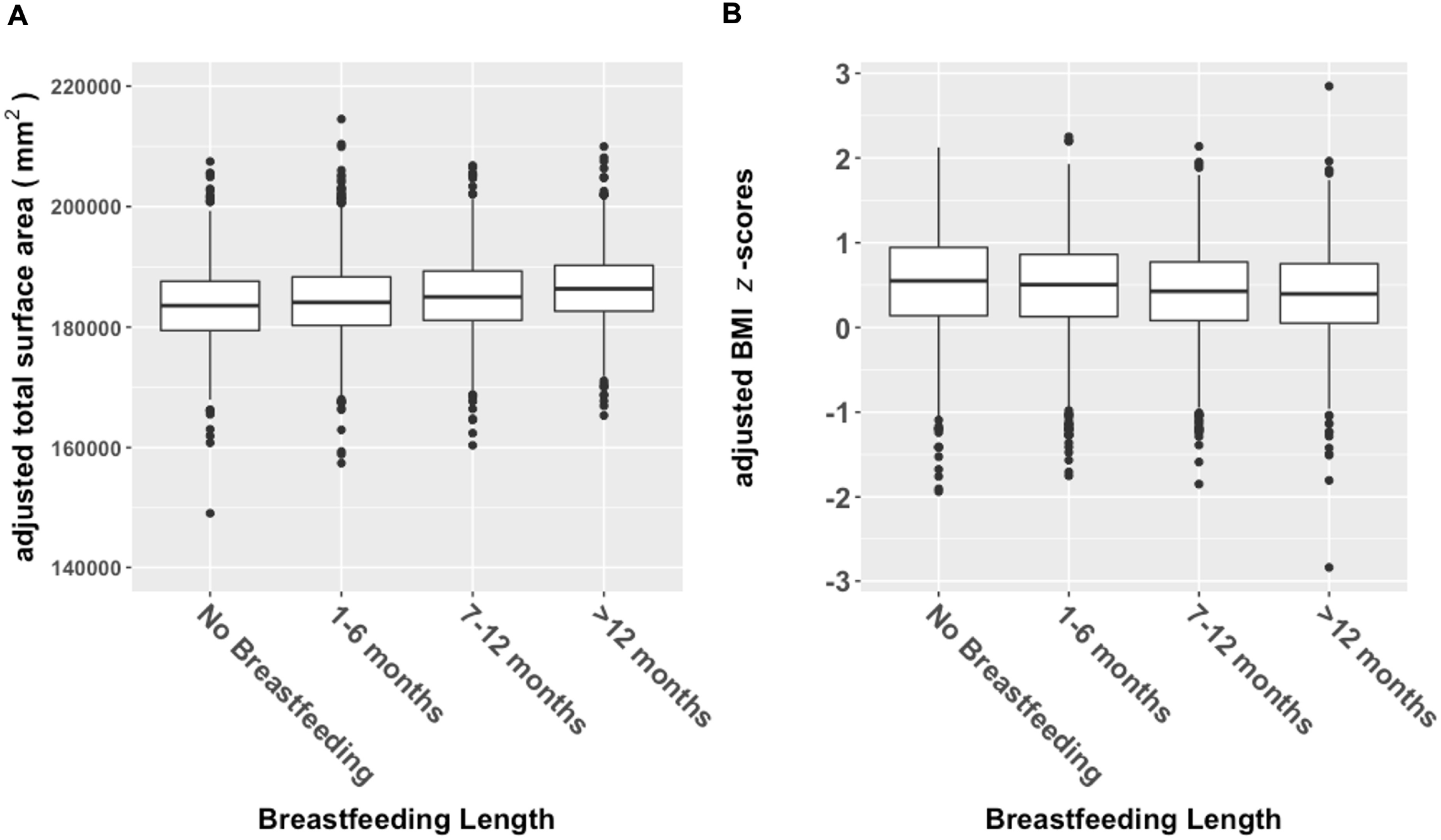
Relationships between breastfeeding length and child brain and adiposity markers. A). Boxplots display distributions of total cortical surface area (adjusted for family ID nested within scanner id, handedness, age, sex, pubertal status, race/ethnicity, family income, parental education, gestational age, maternal health problems during pregnancy, child health problems at birth, maternal age at birth, and maternal alcohol or tobacco use during pregnancy) separated by breastfeeding length category. B). Boxplots display distributions of BMI z-scores (adjusted for family ID nested within site, pubertal status, race/ethnicity, family income, parental education, gestational age, maternal health problems during pregnancy, child health problems at birth, mother’s age at birth, and maternal alcohol or tobacco use during pregnancy) separated by breastfeeding length category.

Similar results were observed from additional analyses where bf duration was dichotomized into any amount of bf vs. no bf (Supplementary Table 1).

### Breastfeeding Length and Child Adiposity Markers

BF duration was negatively associated with adiposity markers (BMI *z*-scores: β (95% CI) = -0.040 (−0.063, -0.016), FDR corrected *P*=0.001, **Figure 1B**; Waist circumference: β (95% CI) = -0.037 (−0.060, - 0.014), FDR corrected *P*=0.002; WHtR: β (95% CI) = -0.040 (−0.064, -0.017), FDR corrected *P*=0.001) (**Table 2**, Supplemental Figure 1B).

Additional analyses examined relationships of adiposity markers with any amount of time bf vs. no bf (Supplementary Table 1) and found similar results.

### Breastfeeding-associated Effects on Brain and Adiposity Markers, modulated by ADI

ADI by bf interactions were not significant for any global brain measurement. Stratified analysis revealed that total cortical SA was significantly correlated with bf length in each ADI tertile (low: (β (95% CI) = 0.050 (0.013, 0.086), FDR corrected *P*=0.030), medium: (β (95% CI) = 0.049 (0.012, 0.086), FDR corrected *P*=0.037), high: (β (95% CI) = 0.054 (0.019, 0.089), FDR corrected *P*=0.006) (**Table 3, Figure 2A**). Total cortical GM volume was significantly correlated with bf length in the low-ADI group (β (95% CI) = 0.023 (0.004, 0.043), FDR corrected *P*=0.042) and high-ADI group (β (95% CI) = 0.030 (0.010, 0.049), FDR corrected *P*=0.006) (**Table 3**, Supplementary Figure 2A).

**Table 3.** Associations between Breastfeeding Length Category and Child Adiposity Markers, Global Brain Measurements, Stratified by Area Development Index (ADI) tertiles.

|  | High ADI |  | Medium ADI |  | Low ADI |  |
| --- | --- | --- | --- | --- | --- | --- |
| Measurement | Std $\beta$<br>(95% CI) | FDR-adjusted<br>*P*-value<sup>a</sup> | Std $\beta$<br>(95% CI) | FDR-adjusted<br>*P*-value | Std $\beta$ (95% CI) | FDR-adjusted<br>*P*-value |
| **Global Brain measurements** |  |  |  |  |  |  |
| Total Cortical Surface area | 0.054 (0.019, 0.089) | **0.006\*\*** | 0.049 (0.012, 0.086) | **0.037\*** | 0.050 (0.013, 0.086) | **0.030\*** |
| Cortical Gray Matter Volume | 0.030 (0.010, 0.049) | **0.006\*\*** | 0.015 (-0.005, 0.034) | 0.199 | 0.023 (0.004, 0.043) | **0.042\*** |
| Subcortical Gray Matter Volume | 0.019 (-0.005, 0.043) | 0.152 | 0.024 (-0.0002, 0.049) | 0.105 | 0.009 (-0.015, 0.033) | 0.601 |
| Cerebral White Matter Volume | 0.011 (-0.026, 0.049) | 0.554 | 0.021 (-0.016, 0.058) | 0.274 | 0.010 (-0.027, 0.047) | 0.601 |
| **Adiposity Markers** |  |  |  |  |  |  |
| BMI *z*-scores | -0.049 (-0.085, -0.001) | **0.046\*** | -0.042 (-0.083, -0.001) | **0.043\*** | -0.027 (-0.068, 0.014) | 0.505 |
| Waist Circumference | -0.042 (-0.084, -0.001) | **0.046\*** | -0.050 (-0.089, -0.011) | **0.019\*** | -0.013 (-0.053, 0.026) | 0.505 |
| Waist-to-Height Ratio | -0.043 (-0.085, -0.001) | **0.046\*** | -0.053 (-0.093, -0.138) | **0.019\*** | -0.017 (-0.058, 0.023) | 0.505 |

<sup>a</sup> \* $P < 0.05$ , \*\* $P < 0.01$

**Figure 2.**
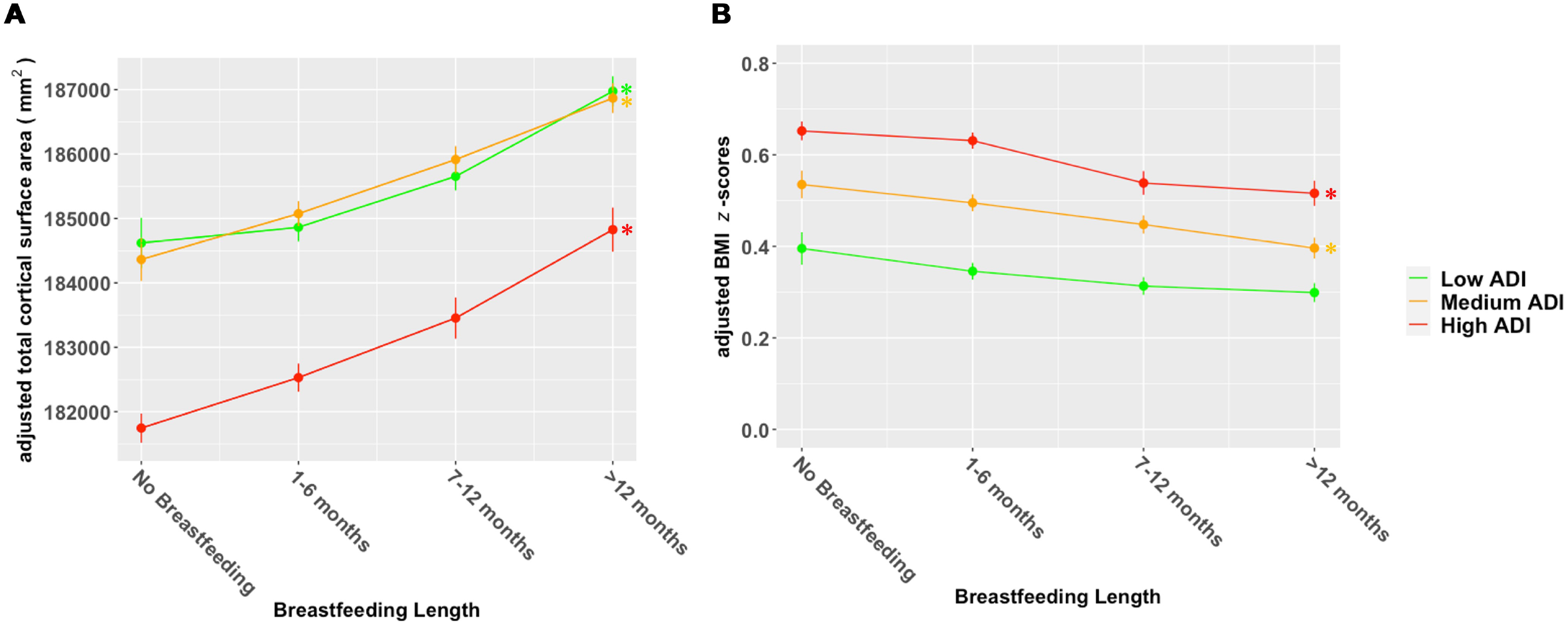
Relationships between breastfeeding length and child brain and adiposity markers in children from neighborhoods with low, medium, and high area deprivation index (ADI). A) Plots display relationship of total cortical surface area with breastfeeding length category, and ADI (in tertiles), with adjustment for family ID nested within site, scanner model, age, sex, pubertal status, race/ethnicity, family income, parental education, handedness, intracranial volume, gestational age, maternal health problems during pregnancy, child health problems at birth, maternal age at birth, and maternal alcohol or tobacco use during pregnancy. All adjustments are made within individual ADI tertiles. B) Plots display relationship of BMI z-scores with breastfeeding length category, and ADI (in tertiles), with adjustment for family ID nested within site, pubertal status, race/ethnicity, family income, parental education, gestational age, maternal health problems during pregnancy, child health problems at birth, maternal age at birth, and maternal alcohol or tobacco use during pregnancy. All adjustments are made within individual ADI tertiles. * P < 0.05

Interaction effects were not observed between ADI and bf on adiposity markers. Stratified analysis showed significant relationships between bf duration and BMI *z*-scores in the high-ADI group (β (95% CI) = -0.049 (−0.085, -0.001), FDR corrected *P*=0.046), and medium-ADI group (β (95% CI) = - 0.042 (−0.083, -0.001), FDR corrected *P*=0.043), but not low-ADI group (β (95% CI) = -0.027 (−0.068, 0.014), FDR corrected *P*=0.505) (**Table 3, Figure 2B**). Similar results were observed across ADI tertiles for waist circumference and WHtR (**Table 3**, Supplementary Figure 2B).

## Discussion

Our findings demonstrated monotonical relationships between bf duration and child global brain measures and adiposity markers even at 9-10 years for the entire study population, with longer bf duration being associated with greater brain measures and smaller adiposity markers. Furthermore, these relationships varied by SEE levels: longer bf duration was associated with lower adiposity in lower SEE children (i.e., high- and medium-ADI). Longer bf duration was associated with greater cortical SA across 3 ADI levels. Our results indicate that long-term benefits of bf on brain and body development in offspring increase as bf length increases and provide scientific evidence for promoting bf, particularly women from low SEEs.

Across the entire study population, the benefits of bf persist into peri-adolescence. Longer bf duration is monotonically related to higher global brain measures, most robustly in cortical SA, cortical and subcortical GM volume. A binary categorization for bf (any amount of bf vs. no bf) yielded similar but less significant effects because data distribution within the bf categories was skewed towards the 1-6 months duration. Our results indicate that beneficial effects of bf to the developing brain increase as bf length increases. Mothers’ bf perceptions are critical to bf duration and reports have shown many mothers find the standard medical recommendations (e.g., exclusive bf for 6 months or longer) overwhelming^32,33^. Our findings indicate that advocacy and education aligned with incremental increase in bf duration could improve bf rates in vulnerable women. With respect to brain benefits, our findings agree with other previous findings where longer bf duration was associated with better neurological and somatic outcomes with durations longer than 6 months having larger effects^34–36^. In line with our findings, positive association of brain GM development with length of bf has been reported across childhood^4^. In contrast to studies reporting improved WM development with longer bf duration, we found a negative correlation^37^. However, prior study results cannot be directly compared to our study as the modalities used to measure WM are not the same. Taken together with published findings across childhood, our findings suggest that bf fosters optimal brain development from infancy to peri-adolescence. These persistent neurodevelopmental benefits may underlie reports of higher IQ, cognitive, and language performances among breastfed children. The protective effects of bf on childhood obesity has been well-established^38,39^. In conjunction with other published studies across childhood, our findings show that longer bf duration is a protective factor for healthy body development well into peri-adolescence. Although specific bf durations were recommended by different organizations, our results indicate that any increment in bf duration provides increased benefits for healthy brain and body growth in children.

Our most prominent finding showed that the benefits of bf, for brain and body development among peri-adolescents, were not uniform across SEE levels. Contrary to prior studies^6,12,40^, we observed that children from lower SEEs (i.e., high- and medium-ADI neighborhoods) showed significantly lower adiposity with longer bf duration. This relationship was not significant in children from low-ADI neighborhood. In parallel, children across all SEE levels demonstrated positive associations between total cortical SA and bf duration. Mixed findings regarding the long-term benefits of bf could be attributed to how various studies accounted for confounders^41–44^. In particular, meta-analyses have shown that the direction and strength of association between bf duration and its long-term benefits changes based on how SEE is measured or modeled in the analysis^45,46^. For example, studies using absolute family income to measure SEE do not account for regional cost-of-living factors. The use of the nationally normed ADI measure allowed for accurate assessment of SEE in a large and diverse population. Additionally, bf duration is strongly correlated with SEE^47^ and therefore many prior studies were underpowered to disentangle the effects of SEE and bf duration on the child’s brain and body development^4^. Leveraging a large and diverse cohort of children in the ABCD study, our study provides critical insight into SEE-independent and SEE-modulated associations between bf and brain and body development in children. Some studies have linked shorter bf duration among low SEE populations to the poor health outcomes to suggest that support longer bf duration in low SEE populations could improve child health outcomes^48–50^. Here, we provided direct evidence showing stronger negative relationships between bf duration and adiposity markers in children from lower SEE levels. Education and support aimed at increasing bf duration for children from lower SEE families, may partly offset their increased obesity risk^51^.

Our study has some limitations. This is an observational and cross-sectional study and therefore cannot establish causal mechanisms underlying these associations. Additionally, while significant, many of the effect sizes measured were small. But this was expected as development across childhood is influenced by multiple factors. We acknowledge that the reported bf durations may be subject to maternal recall bias 9-10 years after its occurrence. Confounders included in this analysis were limited to those available within the ABCD database.

## Conclusions

This is the largest multi-site study investigating the relationships between bf duration and brain structure and adiposity markers in 9-10-year-old children. There were monotonic positive associations of bf duration with global brain measures, and negative associations with adiposity markers. Children from lower SEEs had stronger negative associations between bf duration and adiposity markers. These results suggest that the benefits of bf, on brain and body development, persist into peri-adolescence for the entire study population, and are larger for children from lower SEEs. Interventions and social policies designed to increase overall bf rates and duration has the potential to reduce child health disparities based on SEEs.

## Data Availability

All data produced in the present study are available upon reasonable request to the authors

## Acknowledgements

The authors would like to thank the volunteers who participated in the ABCD Study.

## Author Contributions

Dr. Hsu performed the statistical analysis. Drs. Rajagopalan, Luo and Hsu drafted the manuscript. All authors provided critical review, commentary, and revisions to the manuscript, approved the final manuscript as submitted, and agree to be accountable for all aspects of the work.

## Data Availability Statement

Data for this manuscript were obtained from the ABCD study (https://abcdstudy.org), available from the National Institute of Mental Health Data Archive.

