## Supplemental Materials for "Long-term benefits of breastfeeding on brain and body development among 9–10-year-olds: modulated by socioeconomic environment"

**Supplementary Materials**

**Supplementary Methods**

*Covariates*

Race and/or ethnicity was coded into five categories: White non-Hispanic, Black or African American, Hispanic or Latino, Asian, and Other. Pubertal stage was self-reported by parents using the pubertal development scale, is then reduced to three levels (pre-pubertal, early, mid and above), by combining for all levels mid-puberty and above as one level, in order to pool small samples in late- and post-puberty. Gestational age was derived as 40 weeks for children not born premature, and otherwise by subtracting the number of weeks premature from 40. Maternal health problem(s) during pregnancy was modeled as a binary variable (1 for any problem such as severe nausea or vomiting, heavy bleeding, pre-eclampsia, eclampsia or toxemia, gall bladder attack, proteinurea, rubella, severe anemia, maternal diabetes, urinary tract infections, pregnancy-related high blood pressure, placenta problems, an accident or injury requiring medical care, otherwise 0). Infant’s health problem(s) at birth was modeled as a binary variable (1 for any problem including following, blue at birth, slow heartbeat, did not breath at first, convulsions, jaundice needing treatment, required oxygen, required blood transfusion, Rh incompatibility, otherwise 0). Maternal age at birth was self-reported by the caregiver. Maternal alcohol/tobacco use during pregnancy was coded as a binary variable indicating self-reported use of either substance, either prior to or after knowing of pregnancy.

*Analysis*

Linear mixed effects models were conducted for analysis of correlations of breastfeeding with brain and body, with breastfeeding coded as a binomial variable (No breastfeeding vs. breastfeeding of any amount). Family ID, nested within site, were modeled as random intercepts in models of adiposity markers; and Family ID, nested within scanner ID, were modeled as random intercepts for models of brain measurements. Covariates included age, sex assigned at birth, pubertal status, race/ethnicity, family income, and highest parental education, maternal age, health problems during pregnancy, health problems at birth, gestational age, and maternal alcohol/tobacco use. Age and sex were not included as covariates for models with BMI *z*-score as a dependent variable. Handedness was included in models of brain measurements and intracranial volume was in added in volumetric models.

Supplementary analyses were conducted in R. Linear mixed effects models were fit using the *lme4* package, using Satterthwaite’s method to calculate *P*-values in the *lmerTest* package. Standardized betas were reported, with 95% Wald confidence intervals were calculated based on the local curvature of the likelihood surface. Tests of significance (2-tailed) were corrected for multiple comparisons using the Benjamini-Hochberg false discovery rate (FDR) correction, with *P*<0.05 as the corrected threshold for significance. Cohen’s *d* effect sizes were calculated from least-squares means, model residual standard deviation (SD), and residual degrees-of-freedom using the *emmeans* package.

**Supplementary Results**

*Breastfeeding and Child Global Brain Measurements*

Results for monotonic relationships between breastfeeding duration and other global brain measures were displayed in Supplemental Figure 1A. Breastfeeding of any length (vs. no breastfeeding) was negatively associated with cerebral white matter volume (β (95% CI) = -0.055 (-0.084, -0.026), FDR corrected *P*<0.0001). Although results were not significant, breastfeeding vs. no breastfeeding demonstrated similar data patter as breastfeeding duration in relation to total cortical gray matter volume (β (95% CI) =0.026 (-0.002, 0.054), FDR corrected *P*=0.116) and surface area (β (95% CI) =0.052 (-0.001, 0.104), FDR corrected *P*=0.116). Details see Supplemental Table 1.

*Breastfeeding and Child Adiposity Markers*

Results for monotonic relationships between breastfeeding duration and other adiposity markers were displayed in Supplemental Figure 1B. Breastfeeding of any length (vs. no breastfeeding) was negatively associated with waist circumference (β (95% CI) = -0.069 (-0.128, -0.011), FDR corrected *P*=0.034) and waist-to-height ratio (β (95% CI) = -0.073 (-0.132, -0.013), FDR corrected *P*=0.034).

*Breastfeeding and Child Global Brain Measures and Adiposity Markers in Area Deprivation Index (ADI) tertiles*

Supplemental Figure 2A and 2B displayed relationships between breastfeeding length and other global brain measures and adiposity markers in children from neighborhoods with low, medium, and high ADI.

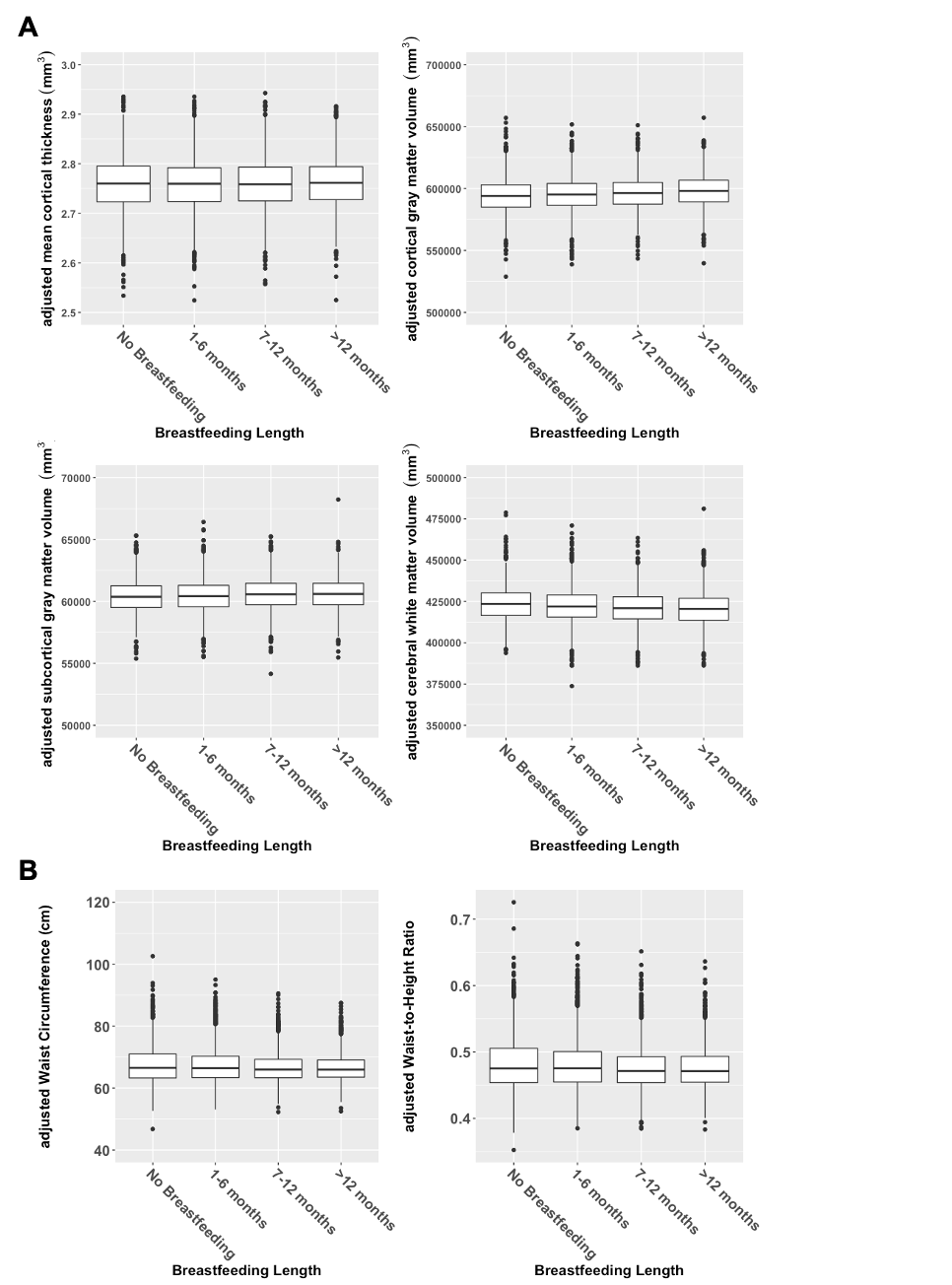

Supplementary Figure 1. Relationships between breastfeeding length and child brain and adiposity markers. A). Boxplots display distributions of mean cortical thickness, total cortical gray matter volume, subcortical gray matter volume, and cerebral white matter volume (adjusting for family ID nested within scanner id, handedness, intracranial volume, age, sex, pubertal status, race/ethnicity, family income, parental education, gestational age, maternal health problems during pregnancy, child health problems at birth, mother’s age at birth, and maternal alcohol or tobacco use during pregnancy) separated by breastfeeding length category. Analysis of breastfeeding and cortical thickness does not adjust for intracranial volume. B). Boxplots display distributions of waist circumference and waist-to-height ratio (adjusting for family ID nested within site, pubertal status, race/ethnicity, family income, parental education, gestational age, maternal health problems during pregnancy, child health problems at birth, mother’s age at birth, and maternal alcohol or tobacco use during pregnancy) separated by breastfeeding length category.

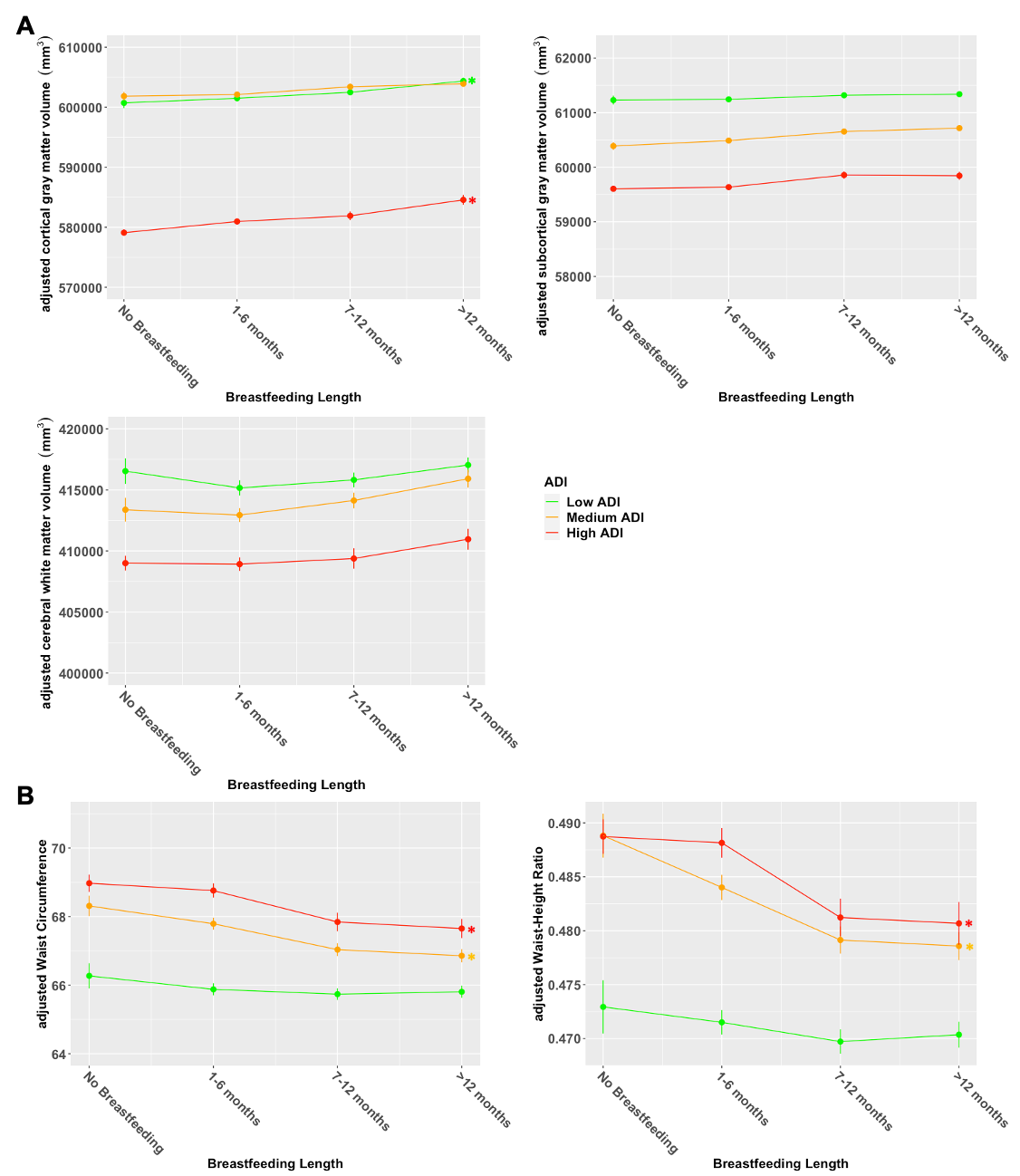
Supplementary Figure 2. Relationships between breastfeeding length and global brain measures and adiposity markers in children from neighborhoods with low, medium, and high area deprivation index (ADI). A) Boxplots display distributions of total cortical gray matter volume, subcortical gray matter volume, and cerebral white matter volume (adjusting for family ID nested within site, scanner model, age, sex, pubertal status, race/ethnicity, family income, parental education, handedness, intracranial volume, gestational age, maternal health problems during pregnancy, child health problems at birth, mother’s age at birth, and maternal alcohol or tobacco use during pregnancy) separated by breastfeeding length category and ADI (in tertiles). B) Boxplots display distributions of waist circumference and waist-to-height ratio (adjusting for family ID nested within site, age, sex, pubertal status, race/ethnicity, family income, parental education, gestational age, maternal health problems during pregnancy, child health problems at birth, mother’s age at birth, and maternal alcohol or tobacco use during pregnancy) separated by breastfeeding length category and ADI (in tertiles).

* FDR Adjusted *P*<0.05

| Supplementary Table 1. Associations between Breasting of any length (vs. no breastfeeding) and Child Adiposity Markers, Global Brain Measurements | | | | | |
| --- | --- | --- | --- | --- | --- |
| **Measurement** | **Std β** | **95% CI** | **Cohen's *d*** | ***P*-value** | **FDR-adjusted *P*-value^a^** |
| **Global Brain measurements** | |  |  |  |  |
| Total Cortical Surface area | 0.052 | (-0.001, 0.104) | 0.101 | 0.054 | 0.116 |
| Mean Cortical Thickness | -0.008 | (-0.063, 0.047) | -0.012 | 0.767 | 0.767 |
| Cortical Gray Matter Volume | 0.026 | (-0.002, 0.054) | 0.082 | 0.07 | 0.116 |
| Subcortical Gray Matter Volume | 0.019 | (-0.015, 0.054) | 0.052 | 0.274 | 0.342 |
| Cerebral White matter Volume | -0.055 | (-0.084, -0.026) | -0.222 | 0.0001 | **0.0009***** |
| **Adiposity Markers** |  |  |  |  |  |
| BMI *z*-scores | -0.057 | (-0.117, 0.003) | -0.087 | 0.064 | 0.064 |
| Waist Circumference | -0.069 | (-0.128, -0.011) | -0.101 | 0.022 | **0.034*** |
| Waist-to-Height Ratio | -0.073 | (-0.132, -0.013) | -0.104 | 0.016 | **0.034*** |

^a^ * *P* < 0.05, *** *P* < 0.001
